# Prognostic Models Predicting Clinical Outcomes in Patients Diagnosed with Visceral Leishmaniasis: A Systematic Review

**DOI:** 10.1101/2024.03.20.24304622

**Authors:** James P Wilson, Forhad Chowdhury, Shermarke Hassan, Eli Harriss, Fabiana Alves, Ahmed Musa, Prabin Dahal, Kasia Stepniewska, Philippe J Guérin

**Affiliations:** Infectious Diseases Data Observatory (IDDO), University of Oxford, Oxford, UK; Centre for Tropical Medicine and Global Health, University of Oxford, Oxford, UK; Bodleian Health Care Libraries, University of Oxford, Oxford, UK; Drugs for Neglected Disease Initiative, Geneva, Switzerland; Institute of Endemic Diseases, University of Khartoum, Khartoum, Sudan

**Keywords:** Leishmaniasis, Visceral, Review, Systematic, Clinical Decision Rules, Prognosis, Prognostic Factors

## Abstract

**Background:** Visceral leishmaniasis (VL) is a neglected tropical disease prevalent in populations affected by poverty and poor nutrition. Without treatment, death is the norm. Prognostic models can steer important management decisions by identifying patients at high-risk of adverse outcomes. We therefore aim to identify, summarise, and appraise the available prognostic models predicting clinical outcomes in VL patients.

**Methods:** We reviewed all published studies that developed, validated, or updated models predicting clinical outcomes in VL patients. Five bibliographic databases were searched from database inception to March 1^st^ 2023 with no language restriction. Screening, data extraction, and risk of bias assessment were performed in duplicate. Findings are presented with tables, figures, and a narrative review.

**Results:** Eight studies, published 2003-21, were identified describing 12 model developments and 19 external validations. All models predicted either in-hospital mortality (n=10 models) or registry-reported mortality (n=2), and were developed in either Brazilian or East African settings (n=9 and n=3 models respectively). Model discrimination (c-statistic) ranged from 0.62-0.92 when evaluated in new data (19 external validations, 10 models). Risk of bias was high for all model developments and validations: no studies presented calibration plots, 11 models were at high risk of overfitting due to small sample sizes, and six models presented risk scores that were inconsistent with reported regression coefficients.

**Conclusion:** With a high risk of bias identified for all models, caution must be exercised when interpreting model predictions and performance measures. Prior to model development or validation, we encourage investigators to review model reporting guidelines. No prognostic models were identified predicting treatment failure or relapse. Furthermore, despite South Asia representing the highest VL burden pre-2010, no models were developed in this population. In the context of the current South Asia elimination programme, these represent important evidence gaps where new model development should be prioritised.

**Registration details:** A protocol for this systematic review has been published (1) and registered (PROSPERO ID: CRD42023417226).

**What is already known on this topic**

- Visceral leishmaniasis (VL) is a neglected tropical disease associated with high mortality, and endemic to regions with constrained resources.
- Identification of high-risk patients is important when prioritising the allocation of limited resources, including inpatient beds, certain VL treatments, and follow-up clinic capacity.
- Risk stratification of VL patients can be performed using prognostic models, however, the range of models, and important model characteristics, have yet to be systematically evaluated.

**What this study adds**

- Following reporting guidelines for systematic reviews of prediction model studies, we present the first comprehensive review of prognostic models that predict clinical outcomes in VL patients.
- We describe 12 prognostic models that all predict mortality in Brazil or East Africa.
- All identified models, including model validations, are assessed at high risk of bias – model predictions and performance measures should be interpreted with caution.

**How this study might affect research, practice or policy**

- This review allows investigators to assess important evidence gaps in the VL prediction model landscape, and identify candidate models for validation or updating using their own patient data.
- Models are identified, summarised, and appraised so that policymakers and healthcare providers can assess model applicability to their own patient population.
- By highlighting limitations in the interpretation of model predictions and performance measures, and to address common sources of bias, we encourage investigators interested in prediction model research to review current guidelines in model reporting, including recently published tools for the calculation of sample sizes and model presentation.

## Introduction

Visceral leishmaniasis (VL), a parasitic infection transmitted between mammalian hosts via the bite of an infected sand fly, disproportionately touches vulnerable people affected by poverty, malnutrition, and forced migration (2). Considered a neglected tropical disease by the World Health Organization (WHO), VL typically presents insidiously with fever, splenomegaly, and weight-loss, and is almost always fatal without effective treatment. The WHO estimates an incidence of 50,000 to 90,000 cases per year, although accurate estimates are obfuscated by incomplete country-level reporting (2,3). Despite progress over the last 20 years, successful treatment remains challenged by drug availability, prolonged treatment courses requiring hospitalisation, and frequent drug side effects (4). Patients with previous treatment failure or immunosuppressive comorbidities such as advanced human immunodeficiency virus (HIV) infection experience particularly high relapse and mortality rates (2,5).

In endemic settings, the accurate identification of high-risk patients is paramount when prioritising the distribution of limited resources; including inpatient beds, treatment regimen, and follow-up intensity. Using prognostic models, typically developed using multivariable regression techniques, the likelihood of a future event occurring can be predicted at the individual patient level (6). Often presented as simplified risk scores, such models abound in the medical literature and support policymakers and healthcare providers in developing treatment guidelines and patient-centred management plans (7,8). Indeed, the number of published prognostic models has steadily surged across range of medical disciplines (8). In infectious diseases alone, recent systematic reviews have identified over 600 prognostic models for COVID-19 (9), 37 models for tuberculosis (10), and 27 models for malaria (11). Despite a burgeoning interest in prediction model research, concerns have been voiced on the conduct and reporting of new models. Biased models can lead to exaggerated estimates of model performance, misleading predictions, and, ultimately, suboptimal and potentially inequitable decision-making (7,8,12).

Prognostic models have been developed and implemented for patients with VL (13,14). The Brazilian Ministry of Health introduced guidelines in 2011 advocating the use of four related risk scores, using a combination of clinical and/or laboratory factors to support whether to admit the patient and treat with liposomal amphotericin B (14–17). Similarly, Médecins Sans Frontières (MSF) Holland in South Sudan have, since 2003, used simple VL risk scores to complement clinical judgement in deciding whether to treat with liposomal amphotericin B, broad-spectrum antibiotics, blood transfusions, and nutritional support (13,18,19). Further models exist (20,21), although the range of models, including their characteristics, comparative performance, and inherent biases, have yet to be systematically described.

We therefore present a systematic review of VL prognostic models, with the aim of identifying, summarising, and appraising prognostic models that predict future clinical outcomes in patients diagnosed with VL. Such a review will allow policymakers to critically assess the implementation of models in treatment guidelines, and healthcare providers to assess the applicability of a particular model to their individual patient, empowering the creation of a patient-centred management plan. Additionally, investigators interested in prediction model research can use this review to identify evidence gaps and assess whether their patient data could be used to develop a new model, or preferably, validate and/or update a pre-existing model (22).

A glossary of key terms relating to model development and evaluation is presented in Box 1.

**Box 1:**
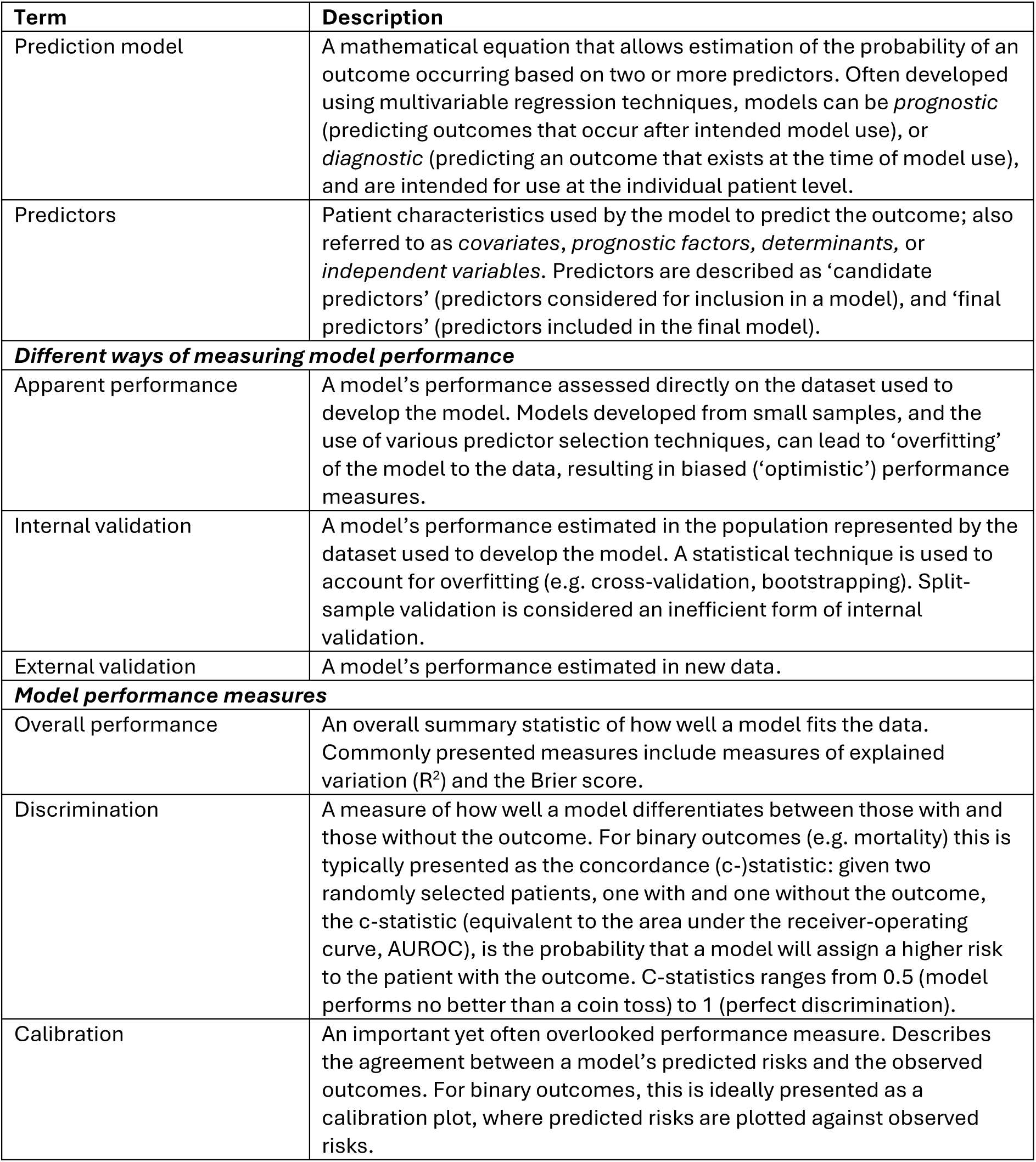
Glossary of key terms relating to prediction model development and evaluation. Adapted from Collins et al 2024 (1) and Moons et al 2015 (2). Collins GS, Dhiman P, Ma J, Schlussel MM, Archer L, Van Calster B, et al. Evaluation of clinical prediction models (part 1): from development to external validation. BMJ. 2024 Jan 8;e074819. Moons KGM, Altman DG, Reitsma JB, Ioannidis JPA, Macaskill P, Steyerberg EW, et al. Transparent Reporting of a multivariable prediction model for Individual Prognosis Or Diagnosis (TRIPOD): Explanation and Elaboration. Ann Intern Med. 2015 Jan 6;162(1):W1–73.

| Term | Description |
| --- | --- |
| Prediction model | A mathematical equation that allows estimation of the probability of an outcome occurring based on two or more predictors. Often developed using multivariable regression techniques, models can be *prognostic* (predicting outcomes that occur after intended model use), or *diagnostic* (predicting an outcome that exists at the time of model use), and are intended for use at the individual patient level. |
| Predictors | Patient characteristics used by the model to predict the outcome; also referred to as *covariates*, *prognostic factors*, *determinants*, or *independent variables*. Predictors are described as ‘candidate predictors’ (predictors considered for inclusion in a model), and ‘final predictors’ (predictors included in the final model). |
| **Different ways of measuring model performance** |  |
| Apparent performance | A model’s performance assessed directly on the dataset used to develop the model. Models developed from small samples, and the use of various predictor selection techniques, can lead to ‘overfitting’ of the model to the data, resulting in biased (‘optimistic’) performance measures. |
| Internal validation | A model’s performance estimated in the population represented by the dataset used to develop the model. A statistical technique is used to account for overfitting (e.g. cross-validation, bootstrapping). Split-sample validation is considered an inefficient form of internal validation. |
| External validation | A model’s performance estimated in new data. |
| **Model performance measures** |  |
| Overall performance | An overall summary statistic of how well a model fits the data. Commonly presented measures include measures of explained variation ($R^2$) and the Brier score. |
| Discrimination | A measure of how well a model differentiates between those with and those without the outcome. For binary outcomes (e.g. mortality) this is typically presented as the concordance (c-)statistic: given two randomly selected patients, one with and one without the outcome, the c-statistic (equivalent to the area under the receiver-operating curve, AUROC), is the probability that a model will assign a higher risk to the patient with the outcome. C-statistics ranges from 0.5 (model performs no better than a coin toss) to 1 (perfect discrimination). |
| Calibration | An important yet often overlooked performance measure. Describes the agreement between a model’s predicted risks and the observed outcomes. For binary outcomes, this is ideally presented as a calibration plot, where predicted risks are plotted against observed risks. |

## Methodology

### Protocol and registration

A protocol for this systematic review has previously been published (1) and registered (PROSPERO registration number: CRD42023417226).

We adhere to Transparent Reporting of Multivariable Prediction Models for Individual Prognosis or Diagnosis: Checklist for Systematic Reviews and Meta-analysis (TRIPOD-SRMA) when reporting this systematic review (23). Data extraction is guided by the Checklist for Critical Appraisal and Data Extraction for Systematic Reviews of Prediction Modelling Studies (CHARMS) (24) and the Prediction Model Risk of Bias Assessment Tool (PROBAST) (7,25). Risk of bias assessment is performed with PROBAST (7,25).

### Eligibility criteria

We follow a Population, Index model, Comparator model, Outcomes, Timing, Setting (PICOTS) approach to frame our review question and define our eligibility criteria (Table 1) (24,26). Studies reporting model development, external validation, updating, or any combination thereof, are considered for inclusion.

**Table 1:** PICOTS approach to frame the research question and define the inclusion criteria. VL: visceral leishmaniasis.

|  |  |
| --- | --- |
| Population | All patients with a confirmed or suspected diagnosis of visceral leishmaniasis as per study authors. |
| Index model | All published prognostic models that develop, validate and/or update (recalibrate/extend) a risk model. |
| Comparator model | Not applicable. |
| Outcomes | Any clinical outcome that occurs after diagnosis. |
| Timing | All prognostic models developed at the time of or following VL diagnosis, and predicting future outcomes. No restriction on the prediction horizon. |
| Setting | No restriction. |

To provide a broad overview of the available prognostic models, we consider all clinical outcomes in patients with a diagnosis of VL and impose no limitation on model setting or prediction horizon (elapsed time period between the intended time of model use and the outcome being predicted).

In accordance with best practice in prediction modelling research (12,25,27), we define a prognostic model as a multivariable model (including two or more predictors) developed with the intention of predicting future outcomes at the individual patient level. Prognostic model studies are distinguished from predictor finding or prognostic factor studies, where the aim is to investigate the effect of a single or group of factors on an outcome of interest (28). We also exclude unpublished studies (including conference abstracts, educational theses), studies that only report diagnostic prediction models, and animal studies.

In addition to studies describing prognostic models, we also identify systematic reviews, meta-analyses, and impact studies of eligible prognostic model and prognostic factor studies. These studies contribute to a narrative review and were not subject to formal data extraction.

### Information sources and search strategy

An information specialist (EH) created the search strategies to retrieve relevant records from the following databases: Ovid Embase; Ovid MEDLINE; the Web of Science Core Collection, SciELO and LILACS. The databases were searched on 1^st^ March 2023 from database inception. The search strategy used text words and relevant indexing terms to retrieve studies describing eligible prognostic models. The Ingui search filter was augmented with an additional search string as described by Geersing et al (29,30), and combined with VL-specific keywords. Google scholar was used to identify any complementary grey literature (full search strategy presented in Supplemental Text 1).

### Study selection

Deduplication and screening of references were performed in Covidence (31). Screening was performed independently by two reviewers (JW, FC); initially at title and abstract level, and subsequently at full-text level. Where discordance existed, a third reviewer (PD), an experienced statistician, was consulted to make the final judgement.

Subsequent forward and backward citation searching were performed to identify records missed by the initial search.

### Data collection process

Study information was captured using a REDCap server hosted at the University of Oxford (32). A data extraction form was created and piloted as per the CHARMS checklist and PROBAST (Supplemental Table 1) (7,24,25). Two reviewers (JW, SH) independently extracted the study information. Where discordance remained after discussion, a final decision was made by a third expert reviewer (PD). Study authors were not contacted in the event of unclear or missing information.

### Risk of bias assessment

Risk of bias was assessed using PROBAST (7,25). Two reviewers (JW, SH) independently assessed each model development (including updating) and external validation by answering 20 signalling questions across four domains (participants, predictors, outcome, and analysis). Responses were used to judge the overall risk of bias as either ‘low’, ‘high’ or ‘unclear’. Any discordance was resolved through discussion.

### Synthesis of results

Information extracted from the identified studies is summarised with a narrative review. Tables and figures facilitate the presentation of important model characteristics. Given our aim is not to compare multiple validations of a single model, no meta-analysis is performed.

### Patient and public involvement

No patients nor members of the public were involved in this systematic review.

### Data sharing and availability

Extracted information is presented in full either in this article and the supplemental material. Our findings will be disseminated through our research group’s website (www.iddo.org/research-themes/visceral-leishmaniasis) and social media channels.

## Results

### Study selection

After deduplication, we identified 2,661 records from the literature search (flow diagram presented in Figure 1). Title and abstract screening identified 61 records for full-text review, of which 11 records were included: eight prognostic model studies (13,14,20,21,33–36) and three systematic reviews (37–39). No impact studies were identified.

**Figure 1:**
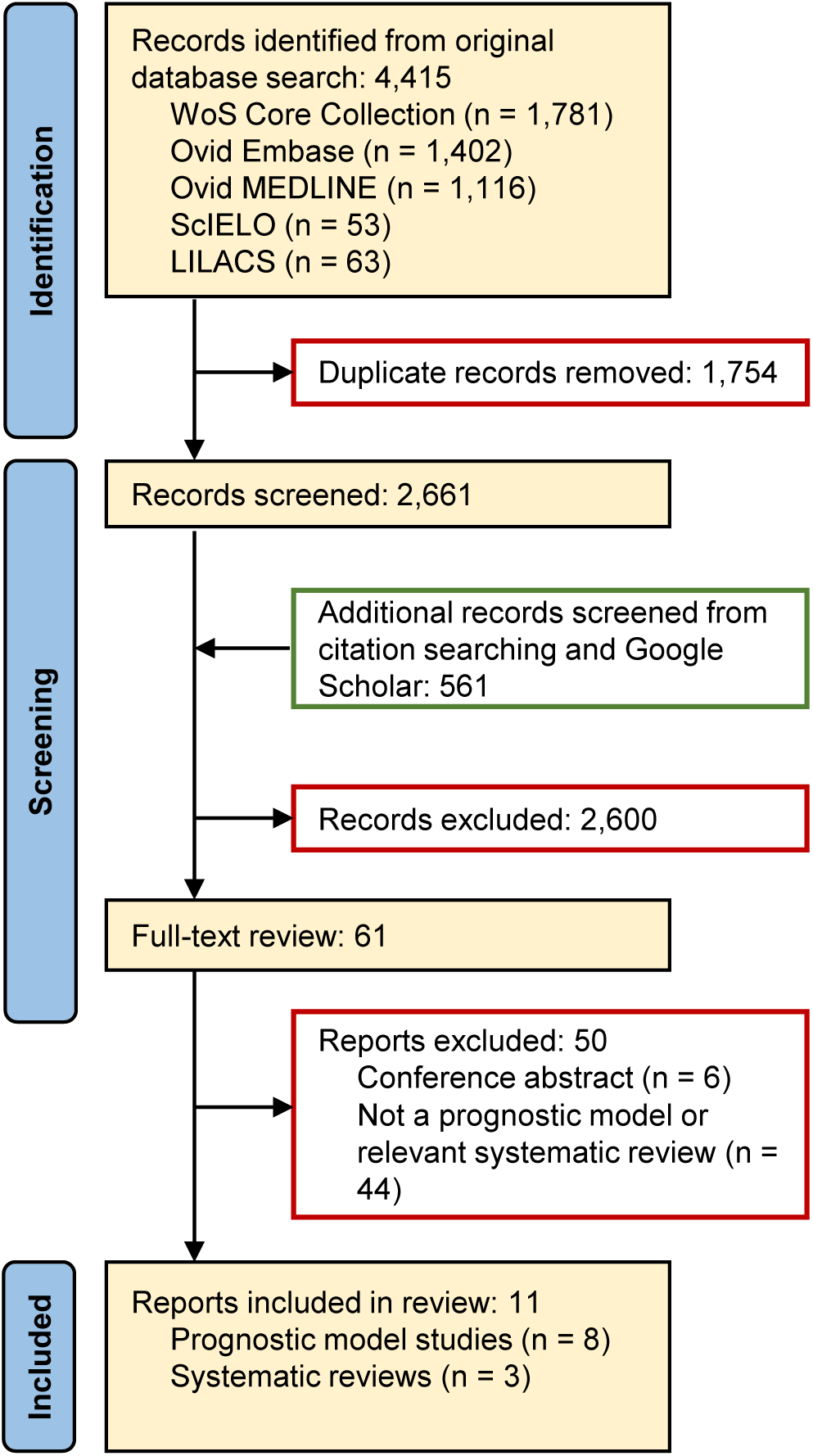
Flow diagram of literature search performed on March 1st 2023, and subsequent record screening.

Of the eight prognostic model studies, four described the development of prognostic models without external validation (20,21,33,34), three described the development of prognostic models with external validation (13,14,35), and one study validated a pre-existing model with subsequent updating (recalibration) (36).

A narrative review of the identified systematic reviews is available in Supplemental Text 2.

### Model characteristics

Important characteristics of the identified models are summarised in Table 2. Each study reported at least one prognostic model, with one study reporting two models (35) and a further study reporting four models (14). Including the updated model (36), a total of 12 models are described, predicting two different outcomes: in-hospital mortality (n=10) (13,14,21,34–36) and registry-reported mortality (n=2) (20,33).

**Table 2:** Key characteristics on the 12 prognostic model developments, ordered by outcome and year published. Each row corresponds to a different model. *Including patients with missing predictor information, excluding patients with missing/excluded outcomes (unless otherwise stated). †Number of predictor parameters (degrees of freedom); for example, a binary or linear predictor is described with 1 parameter; a predictor with 4 categories is described with 3 parameters. Candidate predictors presented in brackets are estimated from incomplete reporting. ‡Number of candidate predictors unclear. Numbers presented are inferred from the study description of extracted information and baseline characteristics. §Number of events not disaggregated by model. ¶Age distribution tabulated by group (not reproduced here). **Sample size excludes both participants with missing predictors and missing/excluded outcomes -, not reported; clin., clinical; conf., confirmed; EPP, events per predictor parameter; excl., excluded; HIV, human immunodeficiency virus; IQR, interquartile range; lab., laboratory; susp., suspected; yrs, years.

| Study | Model described | Data Source | Location<br>Years | Age criteria<br>Diagnostic criteria | Age<br>Spread | % Male | % HIV<br>positive | Events*<br>Sample size (%) | Predictors†<br>Final, Candidate | EPP |
| --- | --- | --- | --- | --- | --- | --- | --- | --- | --- | --- |
| **Outcome: Registry-reported mortality** |  |  |  |  |  |  |  |  |  |  |
| de Araújo 2012 (33) | - | Registry | Brazil (Belo Horizonte)<br>2007-09 | -<br>susp. and conf. | - | - | - | 49<br>376 (13.0%) | 4, 48 | 1.0 |
| Coura-Vital 2014 (20) | - | Registry | Brazil (Nationwide)<br>2007-11 | -<br>conf. | ¶ | 61.7% | 7.0% | 770<br>12,333 (6.2%) | 12, (29) | (26.6) |
| **Outcome: In-hospital mortality** |  |  |  |  |  |  |  |  |  |  |
| Werneck 2003 (21) | - | Case-control | Brazil (Teresina)<br>- (< 2003) | -<br>lab. | Mean: 14.2yrs<br>- | 68.9% | - | 12<br>90 (13.3%) | 4, (15) | (0.8) |
| Sampaio 2010 (34) | - | Retrospective | Brazil (Recife)<br>1996-2006 | < 15 years<br>lab. or clin. | Median: 3.2yrs<br>Range: 4m-13.7yrs | 50.4% | - | 57<br>546 (10.4%) | 6, (15) | (3.8) |
| Costa 2016 (14) | < 2 years, clin. only | Prospective | Brazil (Teresina)<br>2005-08 | < 2 years<br>lab. and clin. | - | - | - | -§<br>314 (-%) | 6, (25) | (0.9) |
|  | < 2 years, clin. + lab. | Prospective | Brazil (Teresina)<br>2005-08 | < 2 years<br>lab. and clin. | - | - | - | -§<br>291 (-%) | 6, (31) | (0.7) |
|  | ≥ 2 years, clin. only | Prospective | Brazil (Teresina)<br>2005-08 | ≥ 2 years<br>lab. and clin. | - | - | - | -§<br>569 (-%) | 9, (27) | (1.6) |
|  | ≥ 2 years, clin. + lab. | Prospective | Brazil (Teresina)<br>2005-08 | ≥ 2 years<br>lab. and clin. | - | - | - | -§<br>538 (-%) | 9, (33) | (1.2) |
| Abongomera 2017 (13) | - | Retrospective | Ethiopia (Abdurafi)<br>2008-13 | -<br>lab. and clin. | Median: 23yrs<br>IQR: 20-28yrs | 95.9% | 19.3% | 99<br>1,686 (5.9%) | 8, 16 | 6.2 |
| Kämink 2017 (35) | < 19 years | Retrospective | South Sudan (Lankien)<br>2013-15 | < 19 years<br>lab. and clin. | ¶ | 54.2% | excl. | 116<br>4,931 (2.4%) | 8, 20 | 5.8 |
|  | ≥ 19 years | Retrospective | South Sudan (Lankien)<br>2013-15 | ≥ 19 years<br>lab. and clin. | ¶ | 56.2% | excl. | 70<br>1,702 (4.1%) | 8, 21 | 3.3 |
| Foinquinos 2021 (36) | Sampaio updating | Retrospective | Brazil (Recife)<br>2008-18 | < 15 years<br>lab. or clin. | - | 48.7% | - | 10<br>156 (6.4%)** | 1, 1 | 10.0 |

Most commonly, studies employed a retrospective cohort design using either hospital records (4 studies, n=5 models) (13,34–36) or national registry data (2 studies, n=2 models) (20,33). One study used a prospective cohort design (1 study, n=4 models) (14) and one study used a case-control design from hospital records (n=1 model) (21). The median number of patients per model development was 542 (range 90–12,333). Over half of all model patients contributed to the largest model from a national registry of confirmed VL cases in Brazil (52.4%, n=12,333) (20).

Most models were developed in Brazil (6 studies, n=9 models) (14,20,21,33,34,36), with the remainder developed in East Africa (one study presenting two models with South Sudanese patients (35), and one study presenting one model with Ethiopian patients (13)). No models were developed in South Asia or the Mediterranean region. Participant age formed the inclusion criteria of eight models (14,34–36), of which five models only included adolescents and younger (14,34–36). No model excluded participants based on sex. Where reported, the median proportion of male participants was 56.2% (range 48.7–95.9%, n=7). Patients living with HIV were either excluded (n=2) (35), not reported (n=8) (14,33,34,36), and where reported, ranged from 7.0–19.3% (n=2) (13,20).

Figure 2 presents the final and candidate predictors identified across model developments, although reporting was inconsistent. The four most common candidate predictors were jaundice (n=12 models), age (n=11), sex (n=10) and bleeding (n=10). Initial VL treatment was included as a candidate predictor in two models, although not included in the final models. Predictors most frequently included in the final models were jaundice (n=11), bleeding (n=8), and age (n=7). No models included sex as a final predictor. Excluding HIV testing, four models did not consider laboratory tests as a predictor (14,20,33). The remaining models (n=8) (13,14,21,34–36) considered and included laboratory tests as predictors in the final model.

**Figure 2:**
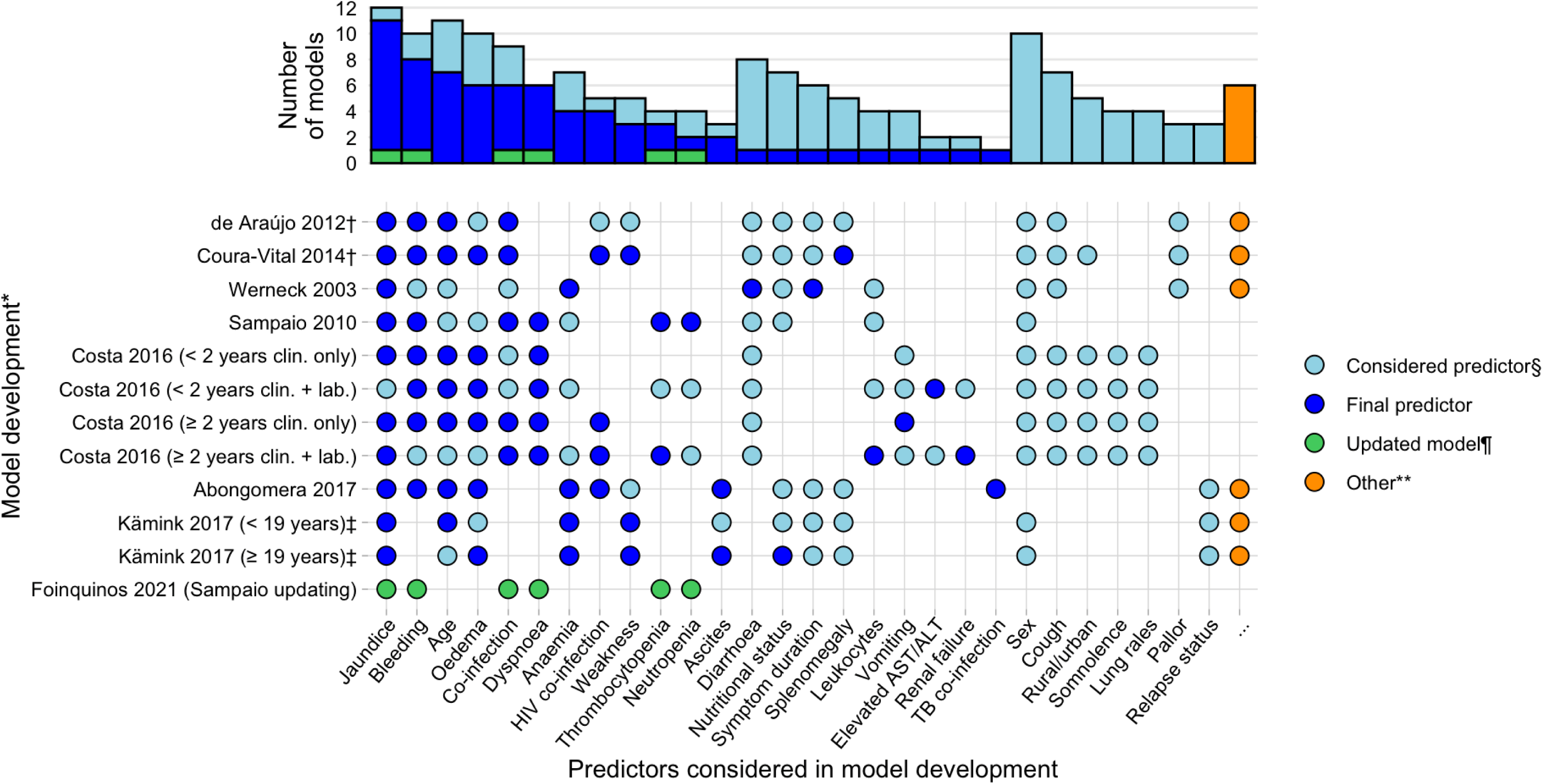
Illustration of the variation of predictors, both considered and included in prognostic model developments. The bar chart presents the number of models incorporating each predictor. To aid comparison, similar predictors have been grouped and renamed. See Supplementary Material for full predictor definitions and groupings. ALT, alanine transaminase; AST, aspartate transaminase; clin., clinical; HIV, human immunodeficiency virus; lab., laboratory; TB, tuberculosis. *Described by author, year, and model name where study presents more than one model †Cough and diarrhoea are combined as a single predictor ‡Oedema and ascites are assessed as a single predictor by [Kämink 2017]. §Considered predictors were assessed, but not included in the final model ¶Predictors in the updated model matched the final predictors in the model being updated [Sampaio 2010] **Predictors assessed in ≤ 2 models and which are not included in the final model: [Werneck 2003]: ‘abdominal distension’, episodes of blood transfusion; [de Araújo 2012]: fever, hepatomegaly, “other clinical manifestations”, initial VL treatment regimen, other VL drugs following initial VL treatment regimen, duration of treatment with antimonials; [Coura-Vital 2014]: fever, hepatomegaly, “other clinical manifestations”, race, education; [Abongomera 2017]: initial VL treatment regimen; [Kämink 2017, both models]: lymphadenopathy.

All 12 models were presented as simplified risk scores, with cut-off thresholds suggested to facilitate clinical decision making. Outcome (i.e. mortality) probabilities corresponding to the risk scores were presented either in tabular format (n=4) (13,20,35), graphically and through a web application (n=4) (14), or not presented (n=4) (21,33,34,36). The full model equation, including model intercept, was reproducible for two models in the development study (20,36), and for a third model in the model updating study (34,36).

Ten of the 12 models underwent one or more external validations; presented either in the development study (n=7 models, 15 validations) (13,14,35,36), or evaluated in separate studies (n=3, 4 validations) (14,36). Two Brazilian studies reported the validation of six models with data collected from the same hospital as model development (developed and validated in either Teresina, state of Piauí (n=5 models) (14,21) or Recife, state of Pernambuco (n=1 model) (36)). Prospectively collected data from Teresina were also used to validate the model developed in Recife (34), and a further model developed using Brazilian national registry data (20). A model developed in Ethiopia (Abdurafi health centre, Abdurafi, Amhara region) was validated using data from a nearby treatment centre (Leishmaniasis Research and Treatment Centre, Gondar, Gondar, Amhara region) (13), and two models developed in South Sudan were validated using retrospectively collected data from both the same treatment centre (Lankien hospital, Jonglei state) and a treatment centre from a neighbouring state in South Sudan (Malakal hospital, Upper Nile state) (35).

Performance measures of discrimination (c-statistic, see Box 1) were presented for all model risk scores. For external validations, the median risk score c-statistic was 0.78 (range 0.62–0.92, n=10 models corresponding to 19 external validations). All studies reported the apparent discriminative performance, with a median risk score c-statistic of 0.86 (range 0.56 – 0.93, n=12 models). A c-statistic was presented for one full model equation (36) and for two model risk scores after accounting for overfitting (internal validation) (13,36). No studies presented overall measures of performance (such as R^2^) or calibration plots. One model’s calibration plot could be reproduced from an internal (split-sample) validation of the risk score (20).

### Risk of bias assessment

Risk of bias assessments for all model developments and external validations, alongside key information on model performance, is presented in Table 3.

**Table 3:**
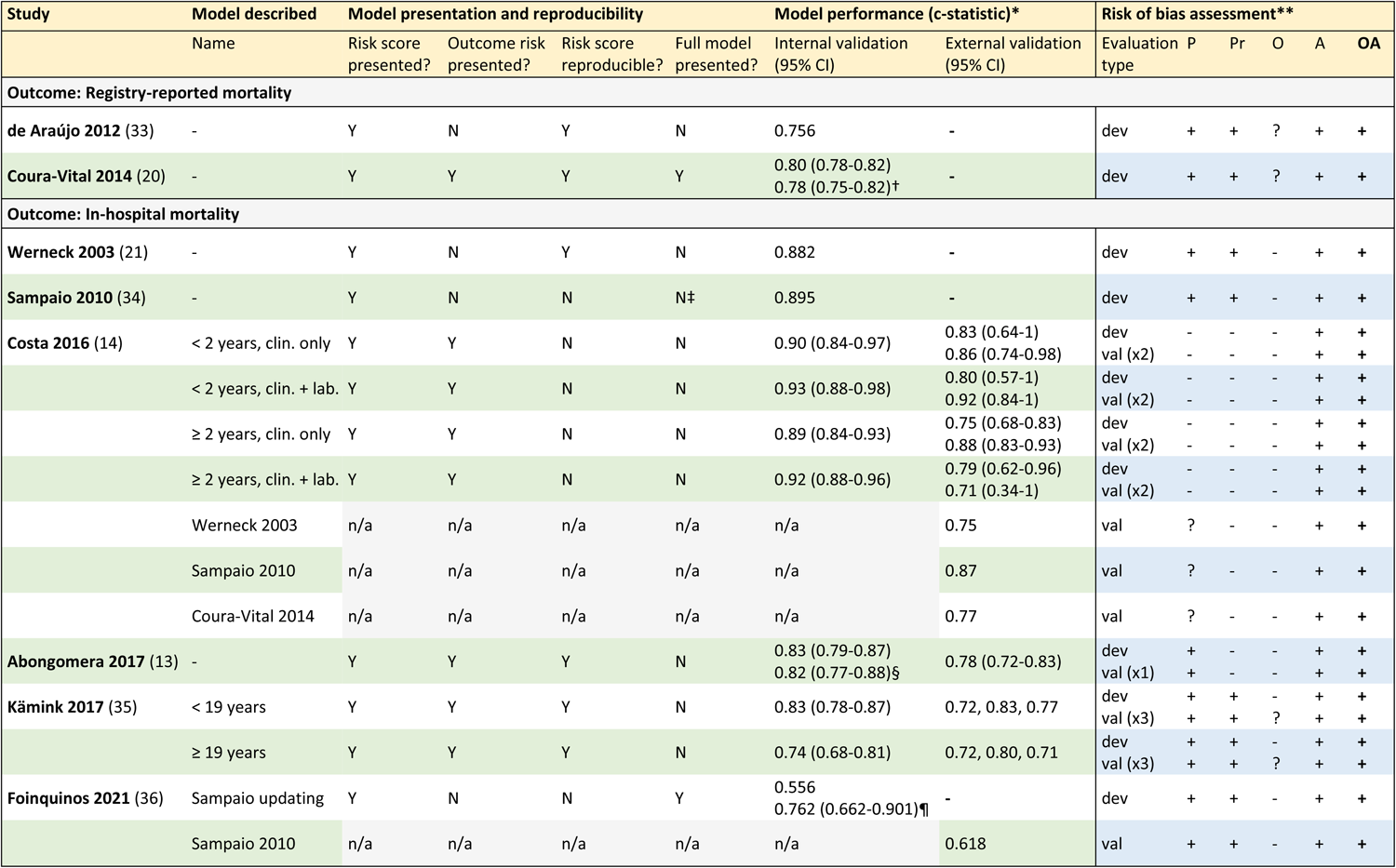
Summary of model presentations and reproducibility for model developments. Performance estimates and risk of bias assessment is presented for both model development (including updating) and external validations. *All internal validation c-statistics relate to the apparent performance of the risk score, i.e. not adjusted for overfitting, unless otherwise stated. 95% confidence intervals are reproduced when reported. Models receiving multiple external validations report multiple c-statistics. the external validations were assessed as having the same risk of bias across all categories and are therefore presented together. †Split-sample (random, 2:1 development:validation). ‡Full regression equation subsequently reported by Foinquinos et al, 2021, when presenting external validation. §Cross-validation (5-fold). ¶Assessing performance of the full model equation. **Assessment of risk of bias is performed separately for model developments, including updating, and external validations. For every model that received more than one external validation +, high risk of bias; -, low risk of bias; ?, unclear risk of bias; -, not reported; c-statistic, concordance-statistic; CI, confidence interval; dev: development; n/a, not applicable for external validations only; N, no; O, outcome; OA, overall assessment; P, participants; Pr, predictors; val, validation; Y, yes.

All 12 model developments were assessed at an overall high risk of bias. Specifically, the model analysis domain was universally assessed at high risk of bias. One model obtained a sufficient sample size (event to predictor parameter ratio > 10), and adequately reported model performance, including calibration (20). All models (excluding the model updating) were developed using a univariable selection stage and did not adjust model predictions to account for optimism due to overfitting. For six models, the presented risk score was not reproducible from the presented regression model coefficients and reported methodology (14,34,36) (calculations reported in Supplemental Text 3).

The outcome domain was assessed at low risk of bias for all model developments, except for two models where bias risk was unclear (20,33). The participants and predictors domains were assessed at high risk of bias for eight (13,20,21,33–36) and seven models (20,21,33–36), respectively.

Specifically, risk of bias was considered high due to differences in predictor definitions and assessments for different patients (n=7) (20,21,33–36) and due to retrospective data collection (n=8) (13,20,21,33–36). One model included predictors that were measured after the likely time of intended model implementation (33).

All extracted information, including characteristics of the external validation datasets, is presented in Supplemental Tables 2-6. This includes participant, predictor, and outcome characteristics (Supplemental Table 2), model analysis, specification, and performance characteristics (Supplemental Table 3), risk score thresholds and interpretations (Supplemental Table 4), risk of bias assessment (Supplemental Table 5), and predictor definitions and categories (Supplemental Table 6).

## Discussion

Across a number of diseases, publications of prognostic models have surged over the last 20 years, driven by an increasing focus on personalised medicine, the need to provide evidence for guideline development, and a growing number of tools available for model development (10,27,40). VL is no exception, with a total of 12 VL prognostic models identified, of which nine were published in the last 10 years (2013–23). We show that all models were developed in Brazil or East Africa, and predict either in-hospital or registry-reported mortality.

When using a prognostic model to predict a VL patient’s mortality risk, for example, in a hospital ward or outpatient clinic, the healthcare provider should be confident that by inputting model predictors (e.g. age, haemoglobin level, clinical signs and symptoms), they receive, as the model output, a reliable estimate of mortality risk. Empowered with this information, the patient can be counselled and shared treatment decisions reached to optimise the available resources and mitigate any increased mortality risk (e.g. allocation of hospital beds and choice of VL treatment). The ability of a model to estimate risk and guide decision making is fundamental: inaccurate risk predictions can lead to suboptimal and inequitable care. Prior to model implementation, VL stakeholders should consider (i) are model risk estimates and performance measures adequately presented, (ii) are they reliable (i.e. subject to bias), and importantly, (iii) how applicable is the model, including its reported performance, to the patient population of interest.

The accuracy of a model’s predictions, where predicted risks and observed outcomes are compared (model calibration, see Box 1), is a fundamental measure of a model’s performance (41,42). However, with calibration only presented adequately for one model (20), we find that VL prognostic model research bucks a broader and concerning trend in prediction model studies: calibration is neglected (9,22,43). Instead, measures of discrimination (the ability of a model to rank patients according to outcome risk, e.g. c-statistic), and classification (where a single risk threshold is assessed, e.g. sensitivity, specificity), are preferentially reported, despite limited clinical utility (7).

All VL model developments were assessed at high risk of bias. Model overfitting, a common source of bias, occurs when the modelling process captures idiosyncratic random variation in the development dataset that is not reflective of the true patient population. Small sample sizes, grouping of continuous predictors, and univariable predictor selection can all introduce bias through overfitting, leading to optimistic performance estimates and exaggerated risk predictions (7,25,44). Bearing this in mind, we demonstrate that all 12 VL prognostic models grouped continuous predictors (such as age and haemoglobin) and included a univariable selection stage in the modelling process. Regarding sample size, the upper limit of events per predictor parameter (EPP) to avoid overfitting is often suggested at 10-20 EPP (7). However, only one model had over 10 EPP (20), with the majority having under 5 EPP. Despite two models reporting model discrimination (c-statistics) adjusted for model overfitting (13,20), no adjusted risk estimates were presented. An important source of bias relates to the accuracy of the derived risk scores: we demonstrate that only half of the risk scores were reproducible from the presented regression coefficients according to the authors’ methodologies, resulting in suboptimal mortality risk estimates.

In four studies, presenting four models, mortality estimates were only reported after dichotomising the risk as ‘high’ or ‘low’ (21,33,34,36). Grouping risk in such a way may guide decision-making in specific settings, but disregards useful information stored in the model, preventing effective use at the bedside or evaluation in new settings. To this end, presentation of the full model equation, including model intercept, is necessary (7). With only two studies presenting the full model equation in the original studies (20,36), investigators wishing to assess the performance of the remaining models in their own data would need to contact the study authors directly.

Visceral leishmaniasis is a complex, heterogeneous disease. The ‘typical’ VL patient and their disease course varies widely between, and often within, endemic areas. This heterogeneity may be explained by a combination of host factors (including age, immunosuppression, malnutrition, and other comorbidities), parasite factors (including genetic variation within and between species), and environmental factors (including vector biology, access to healthcare, and choice of VL treatment) (2,4). The variation in these factors, many of which are unmeasured in routine practice, and surely many additional unknown factors, present prediction challenges when transporting a model from one setting to another. For example, without model updating (recalibration), the application of a model developed in a setting with relatively high inpatient mortality, such as Brazil or a highly HIV-endemic region of East Africa, would likely systematically overestimate mortality in South Asia, a region with relatively low mortality rates (38,39,45). With such variation, and even after disregarding bias introduced through model development, the appealing concept of ‘one model fits all’ becomes untenable. Indeed, the applicability of a model to new settings can only be assessed through external validation (46). Notably, all external validations were performed with datasets from the same country as model development, of which the majority were performed in the same treatment centre using data from a different time period (described as ‘narrow’ or ‘temporal’ validations). Unsurprisingly, all assessed models better discriminate VL patient mortality in the dataset they were developed from compared to the dataset they were evaluated in. This can be explained by true differences in patients, predictors and outcomes not accounted for by the model, and as previously discussed, optimism introduced through overfitting.

Where outcome timing is described, one study reported a third of patients dying within 48 hours of admission (35). A second study reported the average time from treatment initiation to death as 5.5 days and 5.3 days, due to bleeding or co-infection respectively (21). With such a short period between predictor measurement and outcome, it is not surprising that predictors associated with organ failure on the causal pathway to death, such as jaundice, bleeding, co-infection, and dyspnoea, feature prominently in the final model predictors. Interestingly, symptom duration, nutritional status, and relapse status were infrequently included in the final model. Whilst these factors may be causally important; by the time a patient arrives in hospital, symptoms and signs of organ failure are typically more closely associated with death, and therefore preferentially selected for inclusion.

A significant limitation of this review stems from poor reporting of the identified studies. We excluded unpublished manuscripts, including conference abstracts and educational theses, given concerns about quality and difficulty in accessing the original works. Furthermore, in keeping with our prespecified inclusion criteria, this review excluded studies reporting diagnostic models (47,48), models that only evaluated single predictors (49–51), prognostic factor studies (19,52–54), or registered, but unpublished prognostic model studies (for example, NCT05602610). Evaluation of the implementation of prognostic models in current practice and policy was also considered outside the scope of this review.

## Conclusion

In 2015 the TRIPOD Statement was published, providing a 22-item checklist with the aim of facilitating the transparent reporting of prediction model studies (8,27). To reduce model bias and increase model uptake in clinical practice, we encourage investigators considering the development or validation of VL prediction models to consult these guidelines. Additionally, subsequently published resources are now available on how to present models to aid implementation (such as risk scores) (55), and for estimating adequate sample sizes (56,57).

In summary, we present the first systematic review that identifies, summarises, and appraises prognostic models predicting clinical outcomes in patients with VL. We show that no models predict treatment failure or relapse, and no models were developed in South Asia or the Mediterranean region. These represent important gaps in our current knowledge. In the context of the current elimination programme in South Asia, estimating the risk of relapse could help at the population level by identifying infectious and potentially drug-resistant parasite reservoirs (58). Patients identified at high risk of relapse could be selected for more intensive management strategies, including further risk stratification with a parasitological test of cure, and active post-treatment follow-up.

## Data Availability

All data produced in the present work are contained in the manuscript and supplemental material

