## Supplemental Table 1 for "Prognostic Models Predicting Clinical Outcomes in Patients Diagnosed with Visceral Leishmaniasis: A Systematic Review"

*Supplemental Table 1: Information for data extraction and subsequent summary and appraisal. Adapted from the CHARMS Checklist and PROBAST risk of bias tool. VL: visceral leishmaniasis, HIV: human immunodeficiency virus*

| **Domain** | **Key Items** |
| --- | --- |
| Source of data | Source of data (e.g., cohort, case-control, randomised trial participants, registry data, etc.) |
| Participants | Participant eligibility and recruitment method (e.g., location, number of centres, setting, inclusion and exclusion criteria) |
|  | Participant description (age, sex, primary VL or relapse case, co-morbidities including HIV co-infection) |
|  | Details of treatments received |
|  | How VL diagnosis is defined (whether consistent for all participants, using serology and/or microscopy, molecular testing, clinical history and physical signs, etc) |
|  | Study dates |
| Outcome(s) to be predicted | Type of outcome (e.g., single or combined endpoints) |
|  | Definition and method for measurement of outcome (for example, is mortality disease-specific or all-cause, is cure/initial failure/relapse diagnosed based on clinical symptoms and/or diagnostic testing) |
|  | Was the same outcome definition (and method for measurement) used in all patients? |
|  | Time of outcome occurrence or summary of duration of follow-up |
|  | Was the outcome assessed without knowledge of the candidate predictors (i.e., blinded)? |
| Candidate predictors | Number and type of predictors (e.g., demographics, patient history, physical examination, laboratory parameters, HIV status, disease characteristics, etc) |
|  | Definition and method for measurement of candidate predictors (including whether defined and measured in a similar way for all participants) |
|  | Timing of predictor measurement (e.g., at patient presentation, at diagnosis, at treatment initiation or otherwise) |
|  | Handling of predictors in the modelling (e.g., continuous, linear, non-linear transformations or categorised) |
| Sample size | Number of participants and number of outcomes/events |
|  | Events per candidate predictor |
|  | Whether the authors describe a sample size calculation |
| Missing data | Number of participants with any missing value (including predictors and outcomes) |
|  | Number of participants with missing data for each predictor |
|  | Handling of missing data (e.g., complete-case analysis, imputation, or other methods) |
| Model development | Modelling method (e.g., logistic, survival, or other) |
|  | Modelling assumptions satisfied |
|  | Description of participants that were excluded from the analysis with justification |
|  | Method for selection of predictors for inclusion in multivariable modelling (e.g., all candidate predictors, pre-selection based on unadjusted association with the outcome) |
|  | Method for selection of predictors during multivariable modelling (e.g., full model approach, backward or forward selection) and criteria used (e.g., p-value, Akaike information criterion) |
|  | Shrinkage of predictor weights or regression coefficients (e.g., no shrinkage, uniform shrinkage, penalized estimation) |
| Model performance | Calibration (calibration plot, calibration slope, Hosmer-Lemeshow test), discrimination (C-statistic, D-statistic, log-rank), and overall performance measures with confidence intervals |
|  | Classification measures (e.g., sensitivity, specificity, predictive values, net reclassification improvement) and whether a priori cut points were used |
| Model evaluation | Method used for testing model performance: development dataset only (apparent performance, random split of data, resampling methods, e.g., bootstrap or cross-validation, none) or separate external validation |
|  | For external validations; data source and participants to be described as per ‘source of data’ and ‘participants’ domains. Definitions and distributions (including missing data) of outcome and candidate predictors. |
|  | In case of poor external validation, whether model was adjusted or updated (e.g., intercept recalibrated, predictor effects adjusted, or new predictors added) |
| Results | Final and other multivariable models presented, including predictor weights or regression coefficients, intercept, baseline survival, model performance measures (with standard errors or confidence intervals) |
|  | Any alternative presentation of the final prediction models, e.g., sum score, nomogram, score chart, predictions for specific risk subgroups with performance |
|  | Comparison of the definition and distribution of predictors (including missing data) for development and validation datasets |
| Interpretation and Discussion | Study authors’ interpretation of presented models (intended use, clinical utility, etc) |
|  | Study authors’ reported strengths and limitations |
| Miscellaneous | Source of funding / sponsor |
|  | Any declared conflicts of interest |
|  | Methodological guidelines used |
