## Supplemental Table 2 for "Prognostic Models Predicting Clinical Outcomes in Patients Diagnosed with Visceral Leishmaniasis: A Systematic Review"

| **Study details** | | | | | **Predictors** | | **Participants** | | | | **Outcome** | | |
| --- | --- | --- | --- | --- | --- | --- | --- | --- | --- | --- | --- | --- | --- |
| Study +/- model name if > 1 model per study; dev/val/up | | | Data Source | Events/SS (%)^¶^ | **Final**: n (dof)  **Cand**: n (dof), **EPV** | Timing | **Location(s)**  *Study period(s)* | **Sex** (% male)  **Age** (years) (spread) | HIV (% positive) | Eligibility (inclusion and exclusions) | Definition and determination | Timing | LTFU (missing, excluded)  (as % of SS)^¶^ |
| **Outcome: Mortality from registry data** | | | |  |  |  |  |  |  |  |  |  |  |
| **de Araújo 2012** | |  |  |  |  |  |  |  |  |  |  |  |  |
|  |  | **Dev** | Registry^Ꚛ^ | 49/376 (13.0%) | **Final**: 4 (4)  **Cand**: 19 (48)^∑^  **EPV**: 1.02 | ‘time of clinical suspicion’^¥^ | **Brazil:** Belo Horizonte  *2007-09* | **Sex –**  **Age –** | – | (i) Dx: reported to SINAN as ‘suspected’ or ‘confirmed’;  (ii) new case of VL (iii) exclude alt. diag. | Registry reported deaths^Ꚛ^, ‘complemented’ by mortality register | –^µ^ | Death from alt. cause, missing outcome (–) |
| **Coura-Vital 2014** | |  |  |  |  |  |  |  |  |  |  |  |  |
|  |  | **Dev** | Registry^Ꚛ∃^ | **Dev**: 770/12,333 (6.2%)  **Val**: 386/6,168 (6.3%)^#^ | **Final**: 8 (12)  **Cand**: 19 (29)^†^  **EPV**: 26.55 | ‘time of clinical suspicion’ | **Brazil:** Nationwide  *1st Jan 2007-31st Dec 2011* | **Sex**: Dev: 61.7%; Val: 60.7%; **Age**: Dev and Val: by category | **Dev**: 7.0%  **Val**: 6.8% | (i) Dx: reported to SINAN as ‘confirmed’ (ii) exclude alt. diag. | Registry reported deaths^Ꚛ∀^ | – | Death from alt. cause, treatment abandonment, transferred, missing (29.4%) |
| **Outcome: In-hospital mortality** | | | | |  |  |  |  |  |  |  |  |  |
| **Werneck 2003** | |  |  |  |  |  |  |  |  |  |  |  |  |
|  |  | **Dev** | Case-control | 12/90 (13.3%) | **Final**: 4 (4)  **Cand**: 15 (15)^†^  **EPV**: 0.80 | ‘first clinical examination’ | **Brazil**: Infectious Diseases Hospital, Teresina  *Study period not defined* | **Sex**: 68.9%; **Age**: mean: 14.2 (calc) | – | (i) Dx: lab  (ii) notified to hospital epidemiology department | In-hospital mortality from VL (hosp. records) | ‘during treatment | – (–) |
| **Sampaio 2010** | |  |  |  |  |  |  |  |  |  |  |  |  |
|  |  | **Dev** | Retro. cohort | 57/546 (10.4%) | **Final**: 6 (6)  **Cand**: 15 (15)  **EPV**: 3.80 | – | **Brazil**: IMIP, Recife  *May 1996 – June 2006* | **Sex**: 50.4%; **Age**: median – 3.2, (range 4 months – 13.7 years) | – | (i) Dx: lab and/or clin; (ii) < 15 years (iii) exclude alt. diag. | In-hospital mortality (hosp. records) | ‘during treatment’ | –(–) |
| **Costa 2016** | |  |  |  |  |  |  |  |  |  |  |  |  |
|  | < 2 years, clin. | **Dev & Val (x2)** | Pros. cohort | **Dev** –/314  **Val1** –/94  **Val2** –/135 | **Final**: 5 (6)  **Cand**: 24 (25)^†^  **EPV**: (0.9)^‡^ | ‘hospital admission’ | **Brazil**: IDTNP, Teresina  ***Dev****: 1^st^ Sept 2005 – 31^st^ Aug 2008,*  ***Val1****: 1^st^ Sept 2008 – July 31^st^ 2009,*  ***Val2****: 1^st^ Aug 2009 – Nov 31^st^ 2013* | **Sex** –  **Age** – | – | **Dev & Val:** (i) Dx: lab and clin; (ii) age as per model name | **Dev & Val:**  In-hospital mortality (prospective data collection) | **Dev & Val:**  ‘before or during treatment’, ‘discharge or death’ | **Dev & Val:** ‘Presumably’ died from drug toxicities; death after treatment completed (–) |
|  | < 2 years, clin./lab. |  |  | **Dev** –/291  **Val1** –/74  **Val2** –/105 | **Final**: 5 (6)  **Cand**: 30 (31)^†^  **EPV**: (0.7)^‡^ |  |  |  |  |  |  |  |  |
|  | ≥ 2 years, clin. |  |  | **Dev** –/569  **Val1** –/337  **Val2** –/442 | **Final**: 8 (9)  **Cand**: 25 (27)^†^  **EPV**: (1.6)^‡^ |  |  |  |  |  |  |  |  |
|  | ≥ 2 years, clin./lab. |  |  | **Dev** –/538  **Val1** –/70  **Val2** –/104 | **Final**: 8 (9)  **Cand**: 31 (33)^†^  **EPV**: (1.2)^‡^ |  |  |  |  |  |  |  |  |
|  | Werneck 2003 | **Val** |  | – | n/a |  | ***Dev****: 1^st^ Sept 2005 – 31^st^ Aug 2008* |  |  |  |  |  |  |
|  | Sampaio 2010 |  |  |  |  |  |  |  |  |  |  |  |  |
|  | Coura-Vital 2014 |  |  |  |  |  |  |  |  |  |  |  |  |
| **Abongomera 2017** | |  |  |  |  |  |  |  |  |  |  |  |  |
|  |  | **Dev & Val** | Retro. cohort | **Dev**: **99**/1,686 (5.9%)  **Val**: **53**/404 (13.1%)^∃^ | **Final**: 8 (8)  **Cand**: 14 (16)  **EPV**: 6.2 | ‘admission’^€^ | **Ethiopia: Dev**: Abdurafi health centre, Amhere region  *Jan 2008-Dec 2013*  **Val**: LRTC, University of Gondar  *Jan 2011-Dec 2012* | **Sex**: Dev: 95.9%, Val: 97.5%  **Age**: Median (IQR): Dev: 23 (20-28), Val: 25 (20-28) | **Dev**: 19.3%  **Val**: 13.6% | **Dev & Val:**  (i) Dx: as per WHO criteria (clin and/or lab) | **Dev & Val:**  In-hospital mortality (hosp. records) | **Dev & Val:** ‘during admission’ | **Dev:** transferred, defaulted treatment, missing (2.6%)  **Val:** treatment failure, defaulted, missing (5.7%)^∃^ |
| **Kämink 2017** | |  |  |  |  |  |  |  |  |  |  |  |  |
|  | < 19 years | **Dev & Val (x3)** | Retro. cohort | **Dev: 116/**4,931 (2.4%)  **Val** – | **Final**: 4 (8)  **Cand**: 11 (20)  **EPV**: 5.8 | – | **South Sudan:**  **Dev**: Lankien hospital  *July 2013 – June 2015*  **Val1**: Lankian hospital *1999-2002*  **Val2**: Lankian hospital *2002-05*  **Val3**: Malakal hospital *2002-05* | **Sex**: 54.2%; **Age**: by category | n/a | **Dev**:  (i) Dx: clin and lab; (ii) excluding HIV positive  **Val** – | **Dev**: In-hospital mortality (hosp. records)  **Val** – | **Dev:** ‘discharge or death’,  **Val –** | **Dev:** defaulted from treatment, unknown outcome (–, 5.4% for both models combined)  **Val** – |
|  | ≥ 19 years |  |  | **Dev: 70/**1,702 (4.1%)  **Val** – | **Final**: 5 (8)  **Cand**: 11 (21)  **EPV**: 3.3 |  |  | **Sex**: 56.2%; **Age**: by category |  |  |  |  |  |
| **Foinquinos 2021** | |  |  |  |  |  |  |  |  |  |  |  |  |
|  | Sampaio 2010 (validation) | **Val** | Retro. cohort | **10/**156^∃^  (6.4%) | **Final**: 1 (1)  **Cand**: n/a  **EPV**: 10^∞^ | ‘hospital admission’ | **Brazil**: IMIP, Recife  *2008-18* | **Sex**: 48.7%; **Age**: 65.4% < 5 | – | (i) Dx: lab and/or clin; (ii) < 15 years (iii) notified to ‘epidemiology center of hospital’ (iv) exclude alt. diag. | In-hospital mortality (hosp. records) | In-hospital | Death from alt. cause (8.3% with ‘missing records’) |
|  | Sampaio 2020 (updating) | **Up** |  |  |  |  |  |  |  |  |  |  |  |

Table 1: Summary of identified prognostic prediction model studies, ordered by outcome and date published. Extracted information concerning key study characteristics, participants, predictors and outcomes is presented. Each populated row corresponds to a unique model described per study, encompassing model development, validation, updating (recalibration), or any combination thereof.

Abbreviations: –:not reported or disaggregated by model; calc: calculated; EPV: events per variable; Retro: retrospective; LTFU, lost to follow-up; Pros: prospective; n/a: not applicable; CI: confidence interval; Dev: development; SS: sample size; Val: validation; Up: updating

\# Internal validation (split sample, 2:1); not considered a true form of external validation

∑ Including missing data categories

¥ Model development included candidate predictors measured after time of clinical suspicion (other VL drugs following initial VL treatment regimen, duration of treatment with antimonials)

† Number of candidate predictors and/or degrees of freedom unclear. Numbers presented are inferred from the study description of extracted information and baseline characteristics

‡ Number of outcome events not disaggregated by model. Presented EPV is approximated with the overall reported mortality rate of 7.5%

€ Retrospective data collection used a standardised form used in routine clinical practice, and which is presented in the study protocol. Presence of bleeding is collected as a treatment complication and not with admission information

∞Only one regression coefficient being estimated

¶ Unless otherwise stated, the number of events/sample size presented by the study authors *includes* patients with missing predictor information, and *excludes* patients with missing or excluded outcomes

µ ‘Owing to the prolonged incubation period of the disease, data for 2009 were only finalized in March 2010 and, hence, the complete set of information for that year was not available at the time of the study.’

Ꚛ VL registry: Sistema de Informação de Agravos de Notificação (SINAN, The Notifiable Diseases Information System)

∀ Sistema de Informação sobre Mortalidade (SIM, Mortality Information System

∃ Sample size, as presented by authors, excludes both participants with missing predictors and missing/excluded outcomes
