## Supplemental Table 3 for "Prognostic Models Predicting Clinical Outcomes in Patients Diagnosed with Visceral Leishmaniasis: A Systematic Review"

| **Author, year** | | | **Model analysis** |  |  | **Model specification** | | | **Performance^#^** | | | | |
| --- | --- | --- | --- | --- | --- | --- | --- | --- | --- | --- | --- | --- | --- |
| Description, if > 1 model per study, development/validation/  updating | | | Selection of predictors for inclusion in multivariable model (threshold p-value for inclusion) | Multivariable model selection | Handling of particpants with missing predictors (% of SS) | Full regression equation reproducible (including intercept)? | Sum score reproducible using reported methodology? | Sum score presented with corresponding outcome probabilitities? | Apparent  (95% CI) | | Internal  (95% CI) | External  (95% CI) | |
| **Outcome: Mortality from registry data** | | | |  |  |  |  |  |  | | | | |
| **de Araújo 2012** | |  |  |  |  |  |  |  |  | | | | |
|  | - | **Dev** | Univariable logistic regression (p = 0.25), with ‘biologically important’ predictors forced into multivariable stage | Multivariable logistic regression, backwards elimination (p = 0.05) | Missing indicator technique (≥45.2%^‡^) | No | Yes | No | AUC 0.756 | | - | - | |
| **Coura-Vital 2014** | |  |  |  |  |  |  |  |  | | | | |
|  | - | **Dev** | Univariable logistic regression (p = 0.25). ‘Variables […] with collinearity or low frequency were excluded from the multivariate analysis’ | Multivariable logistic regression, backwards elimination (p = 0.05) | Likely complete case^€^ (≥29.8%^‡^) | Yes | Yes | Yes - tabular^∑^ | AUC 0.80 (0.78-0.82) | | AUC 0.78  (0.75-0.82) ^¶^  Calibration tabulated^Ꚛ^ | - | |
| **Outcome: In-hospital mortality** | | | |  |  |  |  |  |  | | | | |
| **Werneck 2003** | |  |  |  |  |  |  |  |  | | | | |
|  | - | **Dev** | Likely univariable logistic regression (p not stated)^£^ | Multivariable logistic regression, likely backwards elimination (p = 0.10) **^£^** | Likely complete case^€^ (-) | No | Yes | No | AUC: 0.882 | | **-** | - | |
| **Sampaio 2010** | |  |  |  |  |  |  |  |  | | | | |
|  | - | **Dev** | Chi-square testing (p = 0.2) | Multivariable logistic regression, backwards elimination (p = 0.05),  interaction terms considered | Likely complete case^€^ (2 with missing weight, 3 with missing FBC; ≥ 0.5%^‡^) | No^¥^ | No | No | AUC: 0.895 | | **-** | - | |
| **Costa 2016** | |  |  |  |  |  |  |  | AUCs | H-L tests |  | AUCs | H-L tests |
|  | < 2 years, clin. | **Dev & Val (x2)** | Likely univariable logistic regression (p = 0.2)^µ^  ‘A variance inflation factor (VIF) was calculated after the regression analysis to assess co-linearity’ | Multivariable logistic regression, backwards elimination (p = 0.05) | Likely complete case^€^ (can infer minimum %s based for the lab/clin models^∞^) | No | No | Yes – graphical^†^, web application | 0.90 (0.84-0.97) | p = 0.52 | - | V1: 0.83 (0.64-1)  V2: 0.86 (0.74-0.98) | V1: p = 0.73  V2: p = 0.61 |
|  | < 2 years, clin./lab. |  |  |  |  |  |  |  | 0.93 (0.88-0.98) | p = 0.66 | - | V1: 0.80 (0.57-1)  V2: 0.92 (0.84-1) | V1: p = 0.69  V2: p = 0.37 |
|  | ≥ 2 years, clin. |  |  |  |  |  |  |  | 0.89 (0.84-0.93) | p = 0.66 | - | V1: 0.75 (0.68-0.83)  V2: 0.88 (0.83-0.93) | V1: p = 0.62  V2: p = 0.54 |
|  | ≥ 2 years, clin./lab. |  |  |  |  |  |  |  | 0.92 (0.88-0.96) | p = 0.93 | - | V1: 0.79 (0.62-0.96)  V2: 0.71 (0.34-1) | V1: p = 0.79  V2: p = 0.43 |
|  | Werneck 2003 | **Val** | n/a | n/a |  | n/a | n/a | n/a | n/a | | n/a | 0.75 | - |
|  | Sampaio 2010 |  |  |  |  |  |  |  |  |  |  | 0.87 | **-** |
|  | Coura-Vital 2014 |  |  |  |  |  |  |  |  |  |  | 0.77 | **-** |
| **Abongomera 2017** | |  |  |  |  |  |  |  |  | | | | |
|  | - | **Dev & Val** | SKJ (Bayesian) methodology. cLHR ≥ 2 or ≤ 0.5 selected for multivariable modelling | Multivariable logistic regression (SKJ approach). aLHR ≥ 1.5 or ≤ 0.67 are selected and model refitted. Procedure repeated until all aLHR ≥ 1.5 or ≤ 0.67 | **Dev**: LHR set to zero for missing data (MCAR assumption)  **Val**: Complete case (18.9% of original n = 525 excluded) | No | Yes | Yes - tabular | AUC 0.83 ( 0.79-0.87) | | AUC 0.82  (0.77-0.88)° | AUC 0.78 (0.72-0.83) | |
| **Kämink 2017** | |  |  |  |  |  |  |  |  | | | AUCs | |
|  | < 19 years | **Dev & Val (x3)** | Univariable logistic regression (p = 0.2) | Multivariable logistic regression, backwards elimination (p not stated) | **Dev**: Complete case (≥ 1.3% for ≥19 model^‡^, ≥0.3% for < 19 model^‡^)  **Val**: Likely complete case^€^ (-) | No | Yes | Yes - tabular^∑^ | 0.83 (0.78-0.87) | | - | Val1: 0.72  Val2: 0.83  Val3: 0.77 | |
|  | ≥ 19 years |  |  |  |  |  |  |  | 0.74 (0.68-0.81) | | **-** | Val1: 0.72  Val2: 0.80  Val3: 0.71 | |
| **Foinquinos 2021** | |  |  |  |  |  |  |  |  | | | | |
|  | Sampaio 2010 (validation) | **Val** | n/a | n/a | Complete case (4.5%) | n/a | n/a | n/a | n/a | | n/a | AUC 0.618  H-L test p = 0.034 (6 groups)  Spiegelhalter test p = 0.007 | |
|  | Sampaio 2020 (updating) | **Up** | Univariable logistic regression; calibration-in-the-large and calibration slope estimated using [Sampaio 2010] model |  |  | Yes | No | No | Sum score: AUC 0.556  Full model: AUC 0.761 (0.622-0.901), Spiegelhalter test p = 0.988 | | **-** | - | |

Table 2: Summary of identified prognostic prediction model studies, ordered by outcome and date published. Extracted information concerning model analysis, specification and performance is presented. Each populated row corresponds to a unique model described per study, encompassing model development, validation, updating (recalibration), or any combination thereof.

Abbreviations: n/a: not applicable; - no information/not reported; CI: confidence interval; Dev: development; Val: validation; Up: updating; [c/a]LHR: crude/adjusted likelihood ratio; MCAR: missing completely at random; SKJ: Spiegelhalter-Knill-Jones; AUC: area under the receiver operating characteristic curve; H-L: Hosmer-Lemeshow; SS: sample size (as presented in Table 1)

\# All performance measures relate to the sum score unless otherwise stated

∑Probability intervals are presented with limits corresponding to the lowest and highest achievable probabilities obtained from the full regression equation, when considering all final predictor permutations corresponding to each total score

¥Full regression equation, including intercept, subsequently specified in Foinquinos 2021 study

†Smoothed lines of best fit are presented showing probability of death by sum score. Methodology unclear

‡Total proportion of missing data across predictors not specified. Percentage represents the greatest amount of missing data per predictor

€Handling of missing data not described, likely complete case

∞For < 2 years subgroup; compared to the clinical only model, the clinical/laboratory model are missing 7.3%, 21.3%, 22.2% for the developmemt, and validation 1 and 2 datasets respectively. Similarly, the for ≥2 years subgroup; the equivalent percentages are 5.4%, 79.2%, 76.5%. Actual numbers not provided, these represent minimum estimates

£ Details unclear

¶Split-sample (random, 2:1 Dev:Val)

°Cross-validation (5-fold)

µ Unclear if univariable stage undertaken once for each model or once for each age group

Ꚛ Calibration of sum score discussed, using two tables it is possible to compare the expected probability range (from development dataset) with observed point probabilities (from validation dataset)
