## Supplemental table 4 for "Prognostic Models Predicting Clinical Outcomes in Patients Diagnosed with Visceral Leishmaniasis: A Systematic Review"

| **Author, year** | | | **Score range** | **Classification measures** | **Rationale for cut-off** | **Suggested risk groups** | **Intended or suggested use of developed or updated model** |
| --- | --- | --- | --- | --- | --- | --- | --- |
| Description, if > 1 model per study, development/validation/  updating | | | (Score cut-off presented +/- presented with classification measures) | For stated cut-off (apparent performance measures unless otherwise stated) |  | Suggested action per risk group | Other comments regarding score interpretation and applicability |
| **Outcome: Mortality from registry data** | | | |  |  |  |  |
| **de Araújo 2012** | |  |  |  |  |  |  |
|  |  | Dev | **0 – 5**  (≥ 1) | Sn: 71.45; Sp: 73.7%; PPV 28.%; NPV 94.5% | ‘For the purposes of comparison’ | **0: -**  **1-5:** should receive specialised clinical management during treatment | ‘The key issue, however, is to define a simple prognosis score that could be applied in basic health units and would allow the early detection of VL cases for redirection to specialized health service. […] The scoring system presented here can be used to identify patients running a higher risk of death from VL at the time of clinical suspicion. Those patients with a score between 1 and 5 should receive specialized clinical management during treatment.’ |
| **Coura-Vital 2014** | |  |  |  |  |  |  |
|  |  | Dev | **0 – 14**  (≥ 4) | Sn: 89.4%; Sp 51.2%  ‘It was observed that the positive predictive values ranged from 8.8% to 37.9% when the case-fatality rate ranged from 5% to 25%. Negative predictive values ranged from 98.9% to 93.6%’ | ‘It is noteworthy that a patient with a score of 4 has approximately 4.5% probability of death from VL, which is relevant from a clinical point of view’ | **0-1: low risk, PPV <1.1%** ‘indicating that outpatient treatment is potentially safe’  **2-3: PPV < 3.2%,** ‘which may indicate treatment during a short stay in the hospital or very close follow-up on an outpatient basis’  **≥ 4: PPV ≥ 3.4 – 5.2%** ‘maximum surveillance and attention’ | ‘According to the clinical point of view and the predictive scoring system proposed here, a patient with a score of 4 or more should have maximum surveillance and attention, because this score presents a risk of death of approximately 4.5% or more. […] Hospitalization should be required for all these groups because specific treatment and measures such as hydration, antipyretics, antibiotics, blood therapy, and nutritional support, should be administered and testing to monitor treatment should be performed ‘  ‘[...] assist decision-making regarding the transfer of the patients to hospitals more capable of handling their condition, admission to the intensive care unit, and adequate support and specific treatment. […]  ‘As with any predictive score, it should not be used in a definitive manner; clinical decisions should remain dependent on clinical judgment’ |
| **Outcome: In-hospital mortality** | | | |  |  |  |  |
| **Werneck 2003** | |  |  |  |  |  |  |
|  |  | Dev | **0 – 4**  (≥ 2) | Sn: 85.7%; Sp: 92.5%; PPV:† 54.6%; NPV:† 98.4% | - | - | ‘[...] to assist in monitoring individuals before and/or after admission, determining allocation of hospital resources, selecting cases for “heroic” treatment, guiding clinical counseling and auditing individual case records.’  ‘The prognostic system based on these four variables performed reasonably well, showing acceptable levels of sensitivity and specificity, indicating that it might be used as a screening procedure in endemic regions of the Third World. The problem lies in what kind of medical decisions should be made after identifying patients at high risk of death.’ |
| **Sampaio 2010** | |  |  |  |  |  |  |
|  |  | Dev | **0 – 9**  (≥ 3) | Sn: 88.7%; Sp: 78.5%; PPV: 32.0%; NPV: 78.5% | ‘the most adequate combination of sensitivity (88.7%), specificity (78.5%), positive predictive value (32.0%), negative predictive value (78.5%) and area under ROC curve (89.5%).’ | - | ‘Isolated risk factors or a combination of them may lead to important clinical decisions such as: platelet or plasma transfusion, use of antibiotics or use of vitamin K. In addition, early identification of unfavorable prognostic factors can facilitate rational allocation of VL cases to be treated as outpatients, admitted to regional hospitals or referred to larger centers with intensive care facilities. Prognostic and severity markers can also be useful for the selection of candidates for clinical trials.’ |
| **Costa 2016** | |  |  |  |  |  |  |
|  | < 2 years, clin. | Dev & Val (x2) | **0 – 9**  (≥ 4) | Sn: 82.6%; Sp: 86.6% | ‘The score for which sensitivity and specificity were the highest was selected’ [maximising Youden index/sum of Sn and Sp] | Discussed. See -> | ‘[by identifying high-risk patients] further referral to higher levels of medical attention such as hospitalization and ICUs could be timelier. Furthermore, the present study may help with the precise allocation of patients to clinical trials as well as to benchmark medical services dedicated to kala-azar […].’  ‘Possible rules of thumb for model use at bedside would be infant referral from primary care to hospital care when the child’s risk of death is greater than the expected 10% mortality. Similarly, referral to ICUs may be considered when the probability of death is >20%, due to the exponential growth of chance of death at this point, which increases to >40% if patients have one more point after mortality reaches the range of 20-30%. Older individuals should be referred to hospitals if their score reaches 5 points. However, if they are older than 40 years and have HIV-1 infection, they should be treated at the hospital with just a single clinical risk factor. If they have ≥6 points, they would be admitted to the ICU. The most important laboratory tests that would signify a medical decision to transfer to ICUs for patients older than 40 years with HIV-1 infection or with clinical complications, are AST >100UK/L in infants, or leukocyte count 1.5mg/dL. Multiple organ failure is suspected in infants with 6 clinical points or 7 clinical-laboratory points and for older people with 7 clinical points, corresponding to the exponential inflection of the curves. On the clinical and laboratory scoring scale for infants, reaching just 5 points indicates a very high risk of death. In these cases, after the available therapies for L. infantum or sepsis, the only alternatives would be organ protection.’ |
|  | < 2 years, clin./lab. |  | **0 – 11**  (≥ 4) | Sn: 95.4%; Sp: 80.7% |  |  |  |
|  | ≥ 2 years, clin. |  | **0 – 13**  (≥ 4) | Sn: 88.4%; Sp: 73.2% |  |  |  |
|  | ≥ 2 years, clin./lab. |  | **0 – 10**  (≥ 3) | Sn: 87.2%; Sp: 82.6% |  |  |  |
|  | Werneck 2003 | Val | n/a | n/a | n/a | n/a | n/a |
|  | Sampaio 2010 |  |  |  |  |  |  |
|  | Coura-Vital 2014 |  |  |  |  |  |  |
| **Abongomera 2017** | |  |  |  |  |  |  |
|  |  | Dev & Val | **-1 – 5**  (-) | Sn, Sp, PPV, NPV presented for all possible score cut-offs | - | **-1: low risk (PPV 1.0%)** ‘This group could be considered in strategies aiming for outpatient/decentralized management and task shifting. Treatment by lower cadres of health professionals could be envisioned. Treatment with SSG could also be safe and appropriate’  **0: intermediate risk (PPV 3.8%)** ‘[…] such patients could be considered for strategies that include a short stay in a health center or hospital followed by outpatient/decentralized management and task shifting’  **1-5: high risk (PPV 10.4-85.7%)** ‘Such patients can be triaged towards a unit with the highest level of care or referred to a better established center. They could be admitted in an intensive care unit and treated by experienced VL clinicians. The following investigations could be done routinely: biochemistry (renal and liver function tests etc.), TB screening (chest radiograph, abdominal ultrasound etc.) and HIV monitoring (CD4 counts). Emergency/resuscitation, safest VL treatment (AmBisome)–AmBisome supplies supported by the WHO and other urgent supportive treatment could be provided: oxygen, blood transfusion, broad spectrum antibiotics and nutritional therapy’ | ‘The score can enable the early detection of VL cases at high risk of death, which can inform operational, clinical management guidelines and VL program management. Busy treatment programs can use this information to organize patient care according to different patient paths, with different levels of care. However, the decisions on how to use the score, and which cut-offs to apply for decision making require careful consideration as this is context-dependent and largely determined by operational factors. We present the diagnostic performance at different cut-offs, allowing the reader to decide on which cut-off to use in their setting. In relatively better resourced settings (eg. non-governmental organization settings), with sufficient human resources, a higher number of patients could receive closer monitoring/more intensive care. In less resourced settings (eg. overwhelmed public hospitals), applying the same cut-offs might not be feasible, and hence a more careful selection of patients for intensive care might be needed.’  ‘Nevertheless, more evidence is needed on the impact of the score when applied for such strategies.’  ‘[...] can inform operational, clinical management guidelines and VL program management. [...] used in clinical research, to standardize patients according to risk groups after inclusion in clinical trials evaluating novel strategies to reduce mortality’ |
| **Kämink 2017** | |  |  |  |  |  |  |
|  | < 19 years | Dev & Val (x3) | **0 – 17**  (≥ 6) | Sn, Sp, PPV, NPV presented for all possible score cut-offs | ‘The optimum threshold can be determined by considering the clinical and operational implications of the sensitivity and specificity of different thresholds’  Threshold chosen to correspond to > 10% mortality risk. | **< 3%: low risk (score of 0-3 in < 19 years, score of 0 in ≥ 19 years):** ‘treated on an ambulatory basis in outpatient treatment centres’  **3-10%: moderate risk (score of 4-5 in < 19 years, score of 1-2 in ≥ 19 years):** ‘treated in an outpatient department, but under close monitoring by experienced clinicians’  **>10%: high risk (score of ≥ 6 in < 19 years, score of ≥ 3 in ≥ 19 years):** ‘should be admitted as inpatients for specialised VL care and intensive monitoring.’ | ‘The optimal threshold [for specialised VL care] will be a compromise between sensitivity and specificity, i.e. a threshold needs to be chosen that includes as many patients with increased risk of dying as possible, whilst maintaining a rational use of scarce resources. Whilst our focus has been to reduce VL mortality by identifying those patients most at risk of dying, the severity scoring system could be a useful tool in the management of patients at lower risk.’  ‘This severity scoring system will be a clinical decision making tool for allocation of VL patients to the appropriate treatment and to minimise the mortality of the VL patients in South Sudan.’  ‘Although one third of deaths occurred within the first 48 hours after admission, which may limit the impact of our severity scoring system on mortality, the predictive ability and simplicity of the system means that it can be easily operationalised and implemented in the field’ |
|  | ≥ 19 years |  | **0 – 9**  (≥ 3) |  |  |  |  |
| **Foinquinos 2021** | |  |  |  |  |  |  |
|  | Sampaio 2010 (validation) | Val | n/a | n/a | n/a | n/a | n/a |
|  | Sampaio 2020 (updating) | Up | **0 – 6**  (≥ 2) | Sn: 30%; Sp: 81.3%; PPV: 10.2%; NPV: 94.2% | ‘A score ≥ 2 was selected as the most adequate predictor of death because it was able to congregate the most satisfactory combination of sensitivity (30%), specificity (81.3%), positive predictive value (10.2%), negative predictive value (94.2%)…’ | - | ‘[...] to predict death in children hospitalised with VL’ [...] ‘applied by health professionals of endemic areas, optimizing the identification of potentially severe cases and the subsequent adoption of earlier interventions such as the early and careful use of antimicrobials, transfusion of blood products, and transfer to intensive care units.’ |

**Supplemental Table 4: Authors’ suggested interpretation of the presented prognostic models. Including sum score range, chosen score thresholds or risk groups (with justification) and apparent classification measures,** Abbreviations: Sn: sensitivity; Sp: specificity; PPV: positive predictive value; NPV: negative predictive value; n/a: not applicable, -: not reported. †Outcome prevalence used to estimate the positive and negative predictive values are is not discussed by the authors.
