## Supplemental text 1 for "Prognostic Models Predicting Clinical Outcomes in Patients Diagnosed with Visceral Leishmaniasis: A Systematic Review"

**Supplemental Text - Search strategy**

Tables 1-5 describe the search terms for each database (Ovid MEDLINE; Ovid Embase, the Web of Science Core Collection, SciELO and LILACS, respectively). All searches performed on 1^st^ March 2023.

Subsequent Google scholar grey literature review: ("visceral leishmaniasis" OR "Kala-azar") AND ("model" OR "prediction" OR "score" OR "prognostic") (run on July 17^th^ 2023)

| Query # | Query terms |
| --- | --- |
| 1 | Leishmaniasis, Visceral/ |
| 2 | ((Leishmaniasis and Visceral) or (Leishmania and infantum) or (Leishmania and donovani) or (Kala and azar)).ti,ab,kw. |
| 3 | 1 or 2 |
| 4 | Validat$.ti,ab. or Predict$.ti. or Rule$.ti,ab. or (Predict$ adj2 (Outcome$ or Risk$ or Model$)).ti,ab. or ((History or Variable$ or Criteria or Scor$ or Characteristic$ or Finding$ or Factor$) adj2 (Predict$ or Model$ or Decision$ or Identif$ or Prognos$)).ti,ab. or (Decision$ adj2 (Model$ or Clinical$)).ti,ab. or (Prognostic adj2 (History or Variable$ or Criteria or Scor$ or Characteristic$ or Finding$ or Factor$ or Model$)).ti,ab. |
| 5 | logistic models/ |
| 6 | decision*.ti,ab |
| 7 | 5 and 6 |
| 8 | 4 or 7 |
| 9 | (Stratification or ROC Curve or Discrimination or Discriminate or c-statistic or c statistic or Area under the curve or AUC or Calibration or Indices or Algorithm or Multivariable).ti,ab. |
| 10 | roc curve/ |
| 11 | 8 or 9 or 10 |
| 12 | 3 and 11 |

Table 1: Search strategy - Ovid MEDLINE

| **Query #** | **Query terms** |
| --- | --- |
| 1 | visceral leishmaniasis/ |
| 2 | ((Leishmaniasis and Visceral) or (Leishmania and infantum) or (Leishmania and donovani) or (Kala and azar)).ti,ab,kw. |
| 3 | 1 or 2 |
| 4 | Validat$.ti,ab. or Predict$.ti. or Rule$.ti,ab. or (Predict$ adj2 (Outcome$ or Risk$ or Model$)).ti,ab. or ((History or Variable$ or Criteria or Scor$ or Characteristic$ or Finding$ or Factor$) adj2 (Predict$ or Model$ or Decision$ or Identif$ or Prognos$)).ti,ab. or (Decision$ adj2 (Model$ or Clinical$)).ti,ab. or (Prognostic adj2 (History or Variable$ or Criteria or Scor$ or Characteristic$ or Finding$ or Factor$ or Model$)).ti,ab. |
| 5 | statistical model/ |
| 6 | decision*.ti,ab. |
| 7 | 5 and 6 |
| 8 | 4 or 7 |
| 9 | (Stratification or ROC Curve or Discrimination or Discriminate or c-statistic or c statistic or Area under the curve or AUC or Calibration or Indices or Algorithm or Multivariable).ti,ab. |
| 10 | receiver operating characteristic/ |
| 11 | 8 or 9 or 10 |
| 12 | 3 and 11 |

Table 2: Search strategy - Ovid Embase

| Query # | Query terms |
| --- | --- |
| 1 | **TS=((Leishmaniasis and Visceral) or (Leishmania and infantum) or (Leishmania and donovani) or (Kala and azar))** |
| 2 | **TS=(Validat$ or Rule$ or (Predict$ near/2 (Outcome$ or Risk$ or Model$)) or ((History or Variable$ or Criteria or Scor$ or Characteristic$ or Finding$ or Factor$) near/2 (Predict$ or Model$ or Decision$ or Identif$ or Prognos$)) or (Decision$ near/2 (Model$ or Clinical$)) or (Prognostic near/2 (History or Variable$ or Criteria or Scor$ or Characteristic$ or Finding$ or Factor$ or Model$)))** |
| 3 | **TI=(Predict$)** |
| 4 | **TS=(Stratification or ROC Curve or Discrimination or Discriminate or c-statistic or c statistic or Area under the curve or AUC or Calibration or Indices or Algorithm or Multivariable)** |
| 5 | **#4 OR #3 OR #2** |
| 6 | **#5 AND #1** |

Table 3: Search strategy - Web of Science Core Collection

| **Query #** | **Query terms** |
| --- | --- |
| 1 | All indexes: ((Leishmaniasis and Visceral) or (Leishmania and infantum) or (Leishmania and donovani) or (Kala and azar)) |
| 2 | All indexes: Validat* or Rule* or (Predict* and (Outcome* or Risk* or Model*)) or ((History or Variable* or Criteria or Scor* or Characteristic* or Finding* or Factor*) and (Predict* or Model* or Decision* or Identif* or Prognos*)) or (Decision* and (Model* or Clinical*)) or (Prognostic and (History or Variable* or Criteria or Scor* or Characteristic* or Finding* or Factor* or Model*))  or Stratification or ROC Curve or Discrimination or Discriminate or c-statistic or c statistic or Area under the curve or AUC or Calibration or Indices or Algorithm or Multivariable |
| 3 | **1 AND 2** |

Table 4: Search strategy – SciELO

| Query # | Query terms |
| --- | --- |
| 1 | (tw:(((Leishmaniasis and Visceral) or (Leishmania and infantum) or (Leishmania and donovani) or (Kala and azar)) )) AND (tw:(Validat* or Rule* or (Predict* and (Outcome* or Risk* or Model*)) or ((History or Variable* or Criteria or Scor* or Characteristic* or Finding* or Factor*) and (Predict* or Model* or Decision* or Identif* or Prognos*)) or (Decision* and (Model* or Clinical*)) or (Prognostic and (History or Variable* or Criteria or Scor* or Characteristic* or Finding* or Factor* or Model*))  or Stratification or ROC Curve or Discrimination or Discriminate or c-statistic or c statistic or Area under the curve or AUC or Calibration or Indices or Algorithm or Multivariable)) |

Table 5: Search strategy - LILACS
