## Supplemental text 2 for "Prognostic Models Predicting Clinical Outcomes in Patients Diagnosed with Visceral Leishmaniasis: A Systematic Review"

**Narrative summary of systematic reviews**

Three systematic reviews were identified that evaluated eligible prognostic model studies or prognostic factor studies (1–3).

The first published review, Cota et al 2011 (1), reviewed studies that described predictors of relapse in patients over the age of 14 years with HIV-VL co-infection, published up to July 2010. The authors identified 1,017 patients enrolled across 18 studies (minimum of 10 patients enrolled per study), of which 17 were conducted in Europe and 6 included multivariable analyses of potential relapse predictors. Risk factors for relapse were identified and included previous VL episodes, baseline CD4+ T-cell count below 100 cells/mL and a lack of increase of CD4+ T-cell count measured at follow-up. A meta-analysis demonstrated a significant association between secondary prophylaxis and reduced relapse risk. Limitations of the review included heterogeneity in study design, treatment and outcome definitions, and small numbers of patients.

The second systematic review, Belo et al 2014 (2), sought to identify a broad range of studies, including educational works, investigating prognostic factors associated with either death, or adverse outcomes independent of death, in Latin America. The literature search was performed between March and September 2011 and 14 studies were identified; all performed in Brazil and presenting multivariable models. Most studies investigated in-hospital death as the outcome of interest, with only two studies using registry data. A total of 5,142 patients were enrolled. Of the predictors amenable to meta-analysis, the following were considered by the authors as ‘potentially strong predictors’; defined as being consistently and significantly associated with adverse outcomes in the majority of studies: jaundice, thrombocytopenia, bleeding, HIV co-infection, diarrhoea, age (both low and high), neutropenia, dyspnoea and bacterial co-infections. The authors recognised the overall poor methodological quality of the studies, limiting the interpretation of their findings.

Lastly, Abongomera et al 2020 (3), performed a systematic review and meta-analysis with the aim of identifying predictors of mortality in patients with VL limited to East Africa. From a search performed in December 2018, 48 studies were identified that enrolled at least 10 patients; including studies reporting both crude and adjusted measures of association. The total number of patients was 150,072. The authors identified 12 prognostic factors (unadjusted) that were evaluated in at least five studies. Of these factors, 10 ‘core’ factors were found to be significantly associated with mortality in the meta-analysis: jaundice, HIV co-infection, tuberculosis co-infection, age (both low and high), oedema, bleeding, anaemia, severe malnutrition, long duration of illness and splenomegaly. As with the other systematic reviews, findings were limited by significant heterogeneity in study design and poor reporting of the included studies.
