## Supplemental text 3 for "Prognostic Models Predicting Clinical Outcomes in Patients Diagnosed with Visceral Leishmaniasis: A Systematic Review"

```
# This is a R script written in base R (R version 4.3.1).
# Date: August 2023.
# Author: James Wilson.

# For each study and model development, we use the presented regression
# coefficients (log odds ratios) to derive the
# simplified score weights according to the study’s methodology

# rc: regression coefficients (log odds ratios)
# aoc: adjusted odds ratios

######################
### de Araujo 2012 ###
######################

rc <- c("other infections" = 1.17,
        "bleeding"         = 1.25,
        "jaundice"         = 2.31,
        "age_60"           = 1.13)

round(rc/min(rc)) # score is reproducible

# Methodology: Barquet et al. referenced

########################
### Coura-Vital 2014 ###
########################

rc <- c("splenomegaly"                = 0.43,
        "edema"                       = 0.57,
        "weakness"                    = 0.50,
        "bleeding"                    = 1.34,
        "jaundice"                    = 0.48,
        "age_0.5"                     = 2.14,
        "age_0.5_1"                   = 1.02,
        "age_19_50"                   = 0.87,
        "age_50_65"                   = 1.34,
        "age_65"                      = 2.25,
        "leishmania-HIV co-infection" = 0.50,
        "bacterial infection"         = 0.63)

round(rc/min(rc)) # score is reproducible

# methodology: Barquet et al. referenced

####################
### Werneck 2003 ###
####################

aor <- c("diarrhea"   = 7.82,
         "jaundice"   = 10.6,
         "fever"      = 8.64,
         "hematocrit" = 15.9)

rc <- log(aor)
round(rc/min(rc)) # score is reproducible

# methodology: Barquet et al. referenced


####################
### Sampaio 2010 ###
####################

aor <- c("jaundice"             = 4.4,
         "dyspnoea"             = 2.8,
         "mucosal bleeding"     = 4.1,
         "asociated infections" = 2.7,
         "platelets"            = 11.7,
         "neutrophils"          = 3.1)

rc <- log(aor)
round(rc/min(rc)) # not reproducible (also see below)

# methodology: Barquet et al. referenced

#########

# correlation with coefficients from Foinquinos 2021
rc <- c("jaundice"                = 1.490908,
        "dyspnoea"                = 1.015233,
        "mucosal bleeding"        = 1.415629,
        "associated infections"   = 1.003439, # denominator
        "platelets"               = 2.458888,
        "neutrophils"             = 1.145403)

exp(rc) # appear consistent
round(rc/min(rc)) # not reproducible

#######################
### Foinquinos 2021 ###
#######################

rc <- c("jaundice"    = 1.490908,
        "dyspnea"     = 1.015233,
        "bleeding"    = 1.415629,
        "infections"  = 1.003439,
        "platelets"   = 2.458888,
        "neutrophils" = 1.145403)

rc_update <- rc * 0.5224832
round((rc_update)/min(rc_update)) # not reproducible

# methodology: Barquet et al (referenced by Sampaio et al)

##################
### Costa 2016 ###
##################

# model: < 2 years, clin.
rc <- c("age < 12"           = 1.4, # groups switched in article
        "bleeding 3-4 sites" = 1.8,
        "bleeding 5-6 sites" = 3.6,
        "edema"              = 1.8,
        "jaundice"           = 0.9)

rc/min(rc) # not reproducible (score includes dyspnoea)

# model: < 2 years, clin./lab.
rc <- c("age < 12"            = 1.1,
        "bleeding 3-4 sites"  = 1.8,
        "bleeding 5-6 sites"  = 4.1,
        "edema"               = 1.4,
        "dyspnea"             = 1.1,
        "AST/ALT"             = 2.5)

rc/min(rc) # not reproducible
2.55/1.05  # max possible score for AST/ALT with rounding error

# model: > or equal to 2 years, clin.
aor <- c("age >40"          = 3.5,
         "2 sites(?exp)"    = 2.3,  # inconsistent with rc, exp(2.3)
         "3 or more sites"  = 10.4, # inconsistent with rc, exp(2.6)
         "HIV"              = 6.1,
         "edema"            = 2.2,
         "jaundice"         = 2.9,
         "dyspnea"          = 2.3,
         "vomiting"         = 2.0,
         "bac infection"    = 2.9)

rc <- c("age >40"          = 1.3,
        "2 sites(?exp)"    = 2.3,  # inconsistent with rc, log(2.3)
        "3 or more sites"  = 2.6,  # inconsistent with rc, log(10.4)
        "HIV"              = 1.8,
        "edema"            = 0.8,
        "jaundice"         = 1.1,
        "dyspnea"          = 0.8,
        "vomiting"         = 0.7,
        "bac infection"    = 1.1)

log(aor)
log(aor)/min(log(aor))
round(log(aor)/min(log(aor))) # not reproducible (different grouping)

# model: > or equal to 2 years, clin./lab.
rc <- c("3 or more sites" = 1.7,
        "hiv"             = 2.2,
        "jaundice"        = 1.0,
        "dyspnea"         = 0.8,
        "bac infection"   = 1.2,
        "leuk"            = 1.3,
        "plat"            = 2.0,
        "renal fail"      = 2.4)

rc/min(rc)
round(rc/min(rc)) # not reproducible

# methodology: Barquet et al. referenced

#######################
### Abongomera 2017 ###
#######################

alhr <-  c(">40"         = 2.22,
           "5-18"        = 0.93,
           "HIV pos"     = 3.04,
           "HIV neg"     = 0.54,
           "Hb < 6.5"    = 2.16,
           "Hb > 6.5"    = 0.67,
           "Bleeding +"  = 3.11,
           "Bleeding -"  = 0.93,
           "Jaundice +"  = 3.21,
           "Jaundice -"  = 0.93,
           "Weakness +"  = 1.64,
           "Weakness -"  = 0.84,
           "Edema -"     = 0.84,
           "Edema +"     = 2.36,
           "Ascites +"   = 5.84,
           "Ascites -"   = 0.92,
           "TB +"        = 1.71,
           "TB -"        = 0.92)

alhr_adj <- alhr[alhr >= 1.5 | alhr < 0.67 ]
round(log(alhr_adj)) # reproducible

# methodology: Speigalhalter-Knill-Jones (Barquet not referenced)

###################
### Kamink 2017 ###
###################

rc <- c("age_2_5"   = 0.98,
        "age_2"     = 2.37,
        "weak_sev"  = 0.92,
        "weak_col"  = 3.13,
        "hb_75_89"  = 0.57,
        "hb_60_74"  = 1.02,
        "hb_60"     = 2.05,
        "jaundice"  = 2.55)

round(rc/min(rc)) # reproducible

rc <- c("weak_sev"    = 0.90,
        "weak_col"    = 1.44,
        "bmi_145_159" = 0.96,
        "bmi_130_144" = 1.02,
        "bmi_13"      = 1.58,
        #"hb_75_89"   = 0.07,
        #"hb_60_74".  = 0.42,
        "hb_60"       = 1.50,
        "oed_asc"     = 1.58,
        "jaundice"    = 1.23)

round(rc/min(rc)) # reproducible

# methodology: Barquet et al. referenced
```
